# Distilling the Knowledge from Large-language Model for Health Event Prediction

**DOI:** 10.1101/2024.06.23.24309365

**Authors:** Sirui Ding, Jiancheng Ye, Xia Hu, Na Zou

## Abstract

Health event prediction is empowered by the rapid and wide application of electronic health records (EHR). In the Intensive Care Unit (ICU), precisely predicting the health related events in advance is essential for providing treatment and intervention to improve the patients outcomes. EHR is a kind of multi-modal data containing clinical text, time series, structured data, etc. Most health event prediction works focus on a single modality, e.g., text or tabular EHR. How to effectively learn from the multi-modal EHR for health event prediction remains a challenge. Inspired by the strong capability in text processing of large language model (LLM), we propose the framework **CKLE** for health event prediction by distilling the knowledge from LLM and learning from multi-modal EHR. There are two challenges of applying LLM in the health event prediction, the first one is most LLM can only handle text data rather than other modalities, e.g., structured data. The second challenge is the privacy issue of health applications requires the LLM to be locally deployed, which may be limited by the computational resource. **CKLE** solves the challenges of LLM scalability and portability in the healthcare domain by distilling the cross-modality knowledge from LLM into the health event predictive model. To fully take advantage of the strong power of LLM, the raw clinical text is refined and augmented with prompt learning. The embedding of clinical text are generated by LLM. To effectively distill the knowledge of LLM into the predictive model, we design a cross-modality knowledge distillation (KD) method. A specially designed training objective will be used for the KD process with the consideration of multiple modality and patient similarity. The KD loss function consists of two parts. The first one is cross-modality contrastive loss function, which models the correlation of different modalities from the same patient. The second one is patient similarity learning loss function to model the correlations between similar patients. The cross-modality knowledge distillation can distill the rich information in clinical text and the knowledge of LLM into the predictive model on structured EHR data. To demonstrate the effectiveness of **CKLE**, we evaluate **CKLE** on two health event prediction tasks in the field of cardiology, heart failure prediction and hypertension prediction. We select the 7125 patients from MIMIC-III dataset and split them into train/validation/test sets. We can achieve a maximum 4.48% improvement in accuracy compared to state-of-the-art predictive model designed for health event prediction. The results demonstrate **CKLE** can surpass the baseline prediction models significantly on both normal and limited label settings. We also conduct the case study on cardiology disease analysis in the heart failure and hypertension prediction. Through the feature importance calculation, we analyse the salient features related to the cardiology disease which corresponds to the medical domain knowledge. The superior performance and interpretability of **CKLE** pave a promising way to leverage the power and knowledge of LLM in the health event prediction in real-world clinical settings.

## Introduction

The rapid adoption of Electronic Health Records (EHR)^1^ has transformed healthcare, offering vast repositories of patient information. In the Intensive Care Unit (ICU), the ability to predict health-related events^2,3^ in advance is paramount for optimizing treatment strategies and improving patient outcomes. EHR is multi-modality data^4^ containing clinical text^5^ (e.g., diagnosis notes) and time series data^6^ (e.g., Electrocardiography (ECG), Electroencephalography EEG), and structured data^7^ (e.g., lab tests). However, existing health event prediction models often focus on a singular modality, such as text^8^ or tabular EHR^9^, presenting a significant challenge in effectively harnessing the entirety of multi-modal EHR data^10^. Health event prediction^11^ is an essential task in the field of medicine. It is the foundation for precision medicine^12^, personalized treatment^13^, etc. With the rapid development of electronic health records (EHR), the data in healthcare becomes more accessible for training the machine learning (ML) models. In the realm of digital health^14^, we can design ML models to precisely predict health event in advance from the EHR data.

Heart failure^15^, a multifaceted clinical syndrome marked by the heart’s compromised ability to pump blood effectively, stands as a formidable challenge within healthcare systems globally. The unpredictable nature of heart failure exacerbations necessitates predictive models that can anticipate events, enabling clinicians to intervene proactively^16^. Hospitalizations and adverse outcomes associated with heart failure place a considerable burden on both patients and healthcare resources^17^. Accurate prediction models offer the potential to enhance patient care, reduce hospitalizations, and optimize treatment strategies^18^. Hypertension, often referred to as the “silent killer,” remains a prevalent cardiovascular condition characterized by elevated blood pressure levels^19^. The insidious nature of hypertension makes it imperative to identify and predict impending events, such as severe complications like strokes and heart attacks^20^. Timely interventions based on accurate predictions can mitigate risks and improve long-term outcomes for individuals living with hypertension^21^. Predictive models tailored to the dynamic nature of blood pressure fluctuations and patient-specific factors are instrumental in shaping personalized care plans. While traditional predictive models have made strides in these domains, the integration of multi-modal data^22,23^ and advanced processing techniques, such as those offered by LLMs^24^, opens new avenues for refined predictions. Predicting events in heart failure and hypertension introduces specific challenges that necessitate a targeted approach. These challenges include the need to assimilate and interpret diverse data modalities within EHRs, ranging from clinical narratives to structured data and temporal trends. Additionally, the intricate interplay of factors contributing to heart failure and hypertension requires models that can capture the complexity of patient health trajectories.

Efforts are put into building health event predictive model on EHR. Some works^25^ use the structured EHR to build the predictive model. Others^26^ use clinical text to predict health events. There are some works^27,28^ that use both structured and text EHR data. Some of them simply use the clinical text as auxiliary information^27^. Others generate the embedding from clinical text and fuse the multi-modal representations to make final predictions. With the wide application of large language model (LLM)^29^, LLM provides a transformative way to build predictive model on multi-modality EHR data^30^. Some previous works are put into how to apply LLM in health event prediction^31^. However, there are still challenges that hinder the landing of LLM applied to health event prediction. Compared to the traditional deep learning models e.g., LSTM, RNN, for text processing, the special characteristics of LLM pose several challenges in the healthcare application. We summarize the challenges from the model and data perspectives as follows.

**LLMs are not scalable and portable for real-world health predictive applications**^**32**^. As we know, directly using the online LLM for inference has privacy issues^33^ and is very expensive^34^. The local model is needed in many real-world clinical scenarios, e.g., hospitals, medical centers, etc^35^. However, the large size of LLM limits its local deployment and the efficiency of inference didn’t meet the real-time requirement of AI healthcare algorithms^36^. **Learning from both clinical text and structured data remains a challenge**. LLM mainly handles the text data, which is only one modality in EHR data. There are other modalities like structured EHR data which could be learned with predictive models, e.g., Transformer. There is a need to effectively learn both modalities in one framework and adapt LLM in the end-to-end training pipeline^37,38^. Meanwhile, the clinical text usually contains much noise^39^, which will mislead the model learning if directly embedded^40^. **How to model the patient similarity in multi-modality learning**. Previous multi-modal methods fuse the embeddings of multi-modal data and cannot mine the latent relations between patients^41^. Learning the patient similarity is inspired by the doctor’s clinical practice which will refer to the past and related patients’ history.

We are motivated to mitigate these challenges by proposing the **CKLE** framework. For the first and second challenge, the **CKLE** framework distills the knowledge from LLM on the cross-modality EHR data. The cross-modality distillation can integrate the LLM’s knowledge into the prediction model without increasing the model complexity. To fully exploit and utilize the knowledge from LLM, we refine and augment the raw clinical text with prompt learning on LLM which can effectively remove the noise. The augmented clinical text from LLM contains less noise and more general textual information with augmentation. To simultaneously mine the patient similarities and the latent cross-modality relations, we design a contrastive loss to model the pairs of patients and each patient’s text-visit pairs. The contributions of this work can be summarized as follows:

- We distill the cross-modal knowledge from LLM to boost the health event prediction and fully exploit the LLM capability by prompting to generate augmented text.
- We design a contrastive distillation loss to learn the multi-modality knowledge from teacher model and similarities between patients at the same time.
- Extensive experiments are conducted on two representative health event prediction tasks to validate the effectiveness of the **CKLE** framework. **CKLE** can achieve competitive prediction performance on the real-world text-rich EHR data.

## Method

### Problem formulation

For each patient, there will be multiple visits *V*_*i*_, where *i* ∈ [1, *m*] indicates the *m* visits to the hospital. Each visit can be represented by the International Classification of Diseases (ICD^42^) codes demonstrating the diagnosis and treatment in the *j*-th visit as *C*_*j*_ = {*c*_1_, *c*_2_, …, *c*_*k*_}, where *k* is a total number of the ICD codes. Additionally, there are attached clinical notes from the doctors for each visit denoted as *N*_*i*_. The dataset and problem can be formulated as follows.

#### Patient visits dataset

The input dataset can be denoted as *D* = *P*_1_, *P*_2_, …, *P*_*n*_ containing *n* patients. For the *i*th patient, *P*_*i*_ = (*V*_*i*_, *N*_*i*_) where each patient has both visits data and text data.

#### Health event prediction

Given the *i*-th patient features of previous *t* − 1 visits 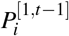, the goal is to train the prediction model *Q*(*θ*) with learnable *θ* parameters, which takes 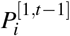 as input and precisely predict the targeted health event *y*_*i*_ at the *t*-th visit of the patient. For heart failure prediction and hypertension prediction, 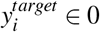 is a binary-class target.

### CKLE framework overview

The overview of the CKLE framework is presented in Figure 1.

**Figure 1.**
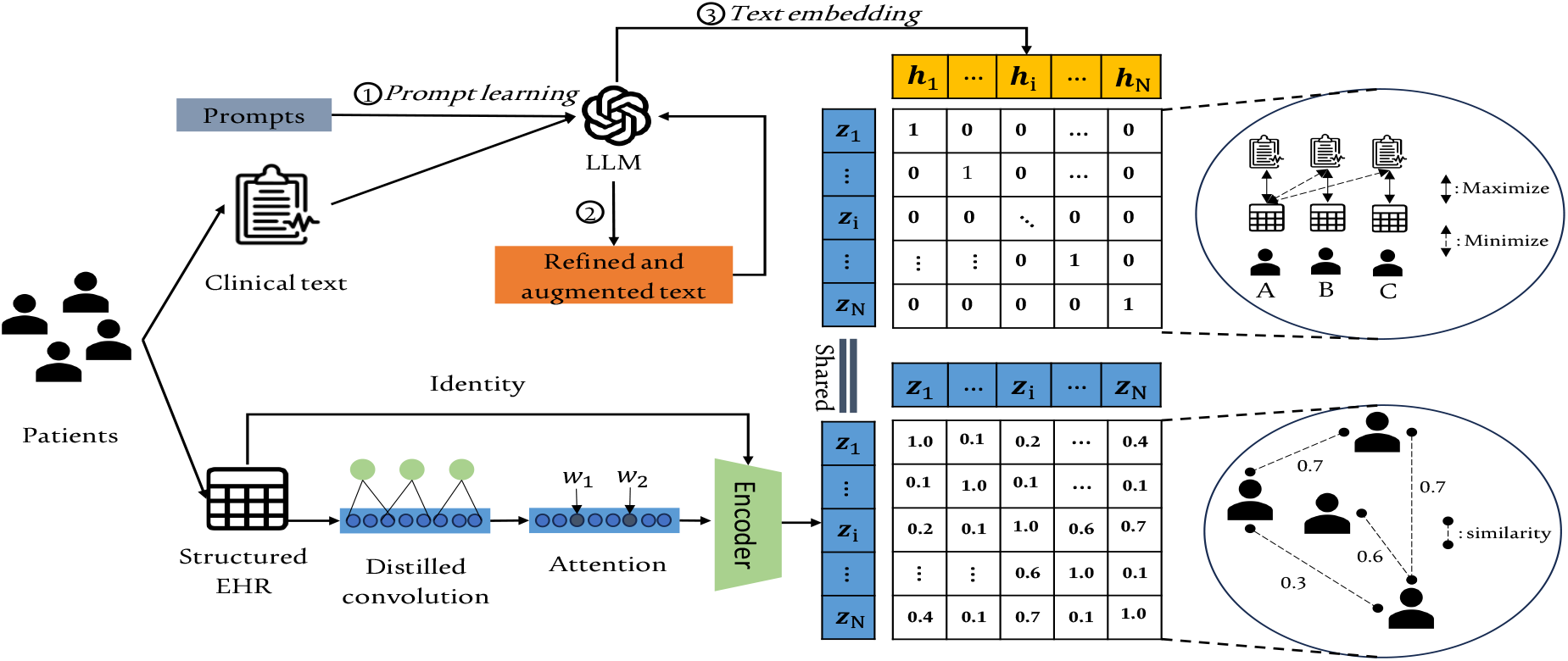
Overview of the CKLE framework.

#### Representation learning from visits data

##### Long-short term feature modelling

We adopt the dilated convolution to learn the long term and short term information from the multiple visits features inspired by^43^. The long-short term feature extraction can be achieved by setting different dilation rate *d*. The dilation convolution layers **dconv** with dilation rate *d* can be represented as follows.

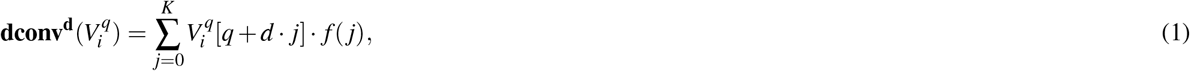

where 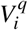 is the *q*-th dimension feature corresponds to *i*-th patient visit *V*_*i*_. The convolution filer with filter size *K* is denoted as *f* (*j*), *j* ∈ [0, *K*]. This illustrates how to learn the representations with a given reception length. Modeling the hidden features at different scale requires multiple convolution with various dilated rate which can be represented as follows.

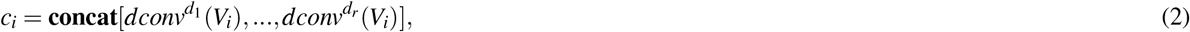

where *c*_*i*_ is the convolution embedding of the *i*-th patient by combining multiple dilated convolution representations with dilated rate from *d*_1_ to *d*_*r*_. *r* is the number of different dilated convolution.

We also employed the feature recalibration module proposed in^43^ to attach suitable attention to different features. The feature recalibration module can be formulated as follows.

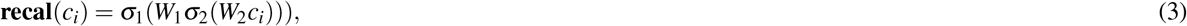

The *c*_*i*_ is hidden representation learnt by dilated convolution. *W*_1_,*W*_2_ are trainable parameters that serves as the features learnable weights. *σ*_1_, *σ*_2_ are activation function which are *Sigmoid, ReLU* respectively. The **recal** weights are then applied to *c*_*i*_ with element-wise multiplying.

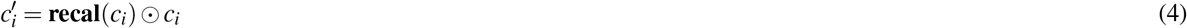

To further improve the representation learning performance, a residual module is applied to remain the original information from patients.

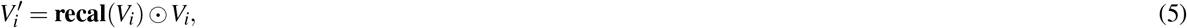

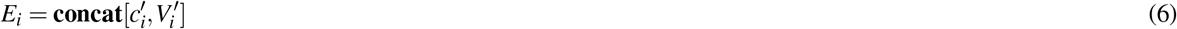

The *E*_*i*_ is the concatenated embedding after the convolution and feature recalibration. The temporal embedding z_i_ will be generated by feeding *E*_*i*_ into a temporal model **temp**, e.g., RNN, GRU.

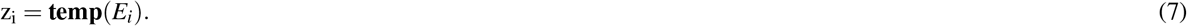

##### Patient similarity modeling

When the doctors are making diagnosis and clinical decisions, they will usually refer to the history of similar patients. Inspired by this process in traditional clinical workflow, we design a contrastive learning module to model the patient similarity which could take advantage of the features from similar patients for health event prediction.

In the conventional contrastive learning setting^44^, the embedding of each patient should be the closet as their own embedding (positive pairs) and farthest to other patients embedding (negative pairs). The target of the contrastive loss is an identity matrix. However, this could not learn the similarity between different patients. So we design a soft target for the contrastive loss to model patient similarity.

We use the ICD codes of each patient as the semantic label **I**_**p**_ of patients. The soft target can be represented as:

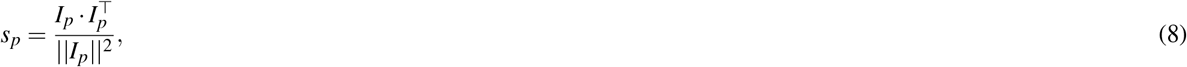

where *s*_*p*_ denotes the similarity between each patient pairs. The logits *y*_*i*_ between patients with batch size |*bs*| are obtained through:

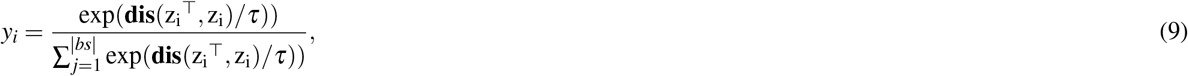

**dis** can be distance computation method which is cosine similarity in this work and *τ* indicates the temperature hyper-parameters. Similarly, the target of patient-patient pairs can be calculated with *s*_*p*_ as:

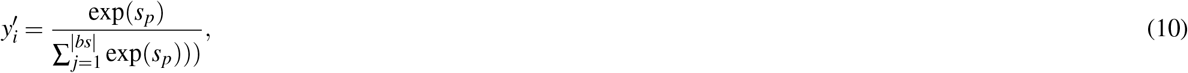

Thus the loss function *L*_psim_ for patient similarity modeling can be represented as:

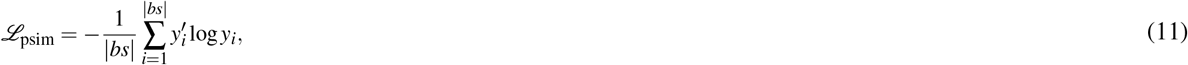

#### Exploit medical knowledge from LLM

##### Text augmentation with prompt learning

The raw clinical text contains unignorable noise and redundant information, which indicates the spelling errors, abbreviations, non-standard terminology, and extraneous information that hinder data extraction and analysis. To remove the unrelated information and increase the generalizability of text, we propose to achieve the text augmentation with prompt learning on LLM by refining the raw text and generating additional synthetic text data based on existing clinical text. For each patient *i*, we have raw notes *N*_*i*_. The *LLM* with frozen knowledgeable parameters are exploited to augment and polish the raw text with prompt learning. The prompt *E* input to the LLM are designed as *“Refine the following clinical text without changing its meanings:”*. Then the prompt will be attached with each patient’s clinical notes *N*_*i*_. The augmented and refined clinical text can be generated as follows.

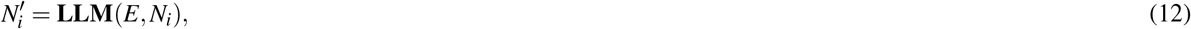

where 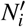 is the augmented and refined notes.

##### Cross-modality distillation from LLM

Each patient has clinical text and tabular data, we obtain the embedding z_i_ of tabular data through the representation learning on multiple visits. To further take advantage of the strong power of LLM, we generate the embedding h_*i*_ of clinical text from the LLM as follows.

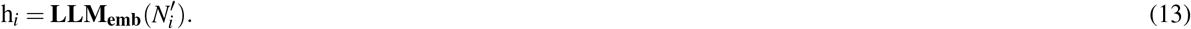

To exploit the rich information from multi-modality, a cross-modality distillation strategy is designed to transfer the knowledge from LLM to the health prediction model. The **LLM** serves as the teacher model with frozen parameters and the prediction model **temp** is the student model with parameters *θ*. The z_*i*_, h_*i*_ are structured EHR and clinical text embedding generated by **LLM** and **temp** respectively.

As each patient has structured EHR and clinical text, the z_*i*_, h_*i*_ from the same patient *P*_*i*_ are highly related. On the other hand, the EHR and clinical text from different patients shares little common information for which the embeddings should have larger distance. Inspired by this domain knowledge, the distillation objective *ℒ*_cmkd_ is designed to learn the contrastive relations^45^ between EHR-text pairs formulated as:

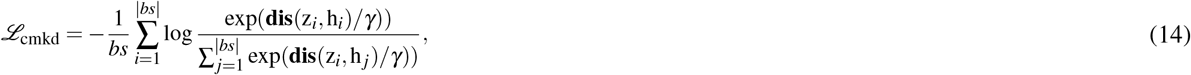

where *γ* is the temperature hyper-parameters.

### Training for CKLE

The final prediction of the health event can be represented as:

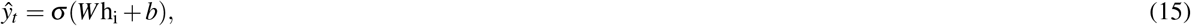

where *W, b* are learnable parameters and *σ* is the activation function, like *sigmoid*. The overall loss function *ℒ* of **CKLE** can be represented by combining these objectives together.

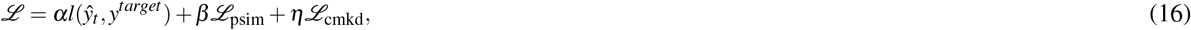

where *l* is the conventional prediction loss function, e.g., cross entropy. *y*^*target*^ is the ground truth label. *α, β, η* are hyper-parameters to control the ratio of different objectives.

### Experimental Setup

#### Dataset and tasks

The well-known medical EHR dataset MIMIC-III is used for the experiments. We filter out to get patients with both clinical text and the corresponding structured data. Inspired by^27^, the patients with multiple visits will be used for the health event prediction. Each patient’s previous visits will be used to predict the last visit. Due to each patient has a variable number of previous visits, we will align them with the largest number of the previous visits among the patients. Two tasks related to cardiology diseases are selected as the representative health event prediction tasks in this work. Details of each task will be introduced as follows.

- Hypertension prediction: This task is a binary classification to predict the hypertension of the patient’s next hospital visit.
- Heart failure prediction: This task is a binary classification, which predicts whether the heart failure will happen to the patient in the next hospital visit.

#### Baselines

- RETAIN: A widely used interpretable healthcare prediction framework with the reverse attention module proposed by Choid et al^46^.
- AdaCare: Ma et al.^43^ designed the AdaCare framework for representation learning on EHR data by modeling the short and long term features and provide explainability with competitive performance.
- Dipole: An interpretable prediction framework based on bi-directional RNN is proposed by Ma et al.^47^. Dipole can memorize the long-term history information and provide clinical meaningful interpretation.
- CGL: This is a specialized health event prediction framework^27^ with text-rich EHR data. CGL use graph learning to learn the patient similarities for event prediction.
- Chet: It is proposed by Lu et al^25^ to use dynamic disease graph to learn the temporal variation of diseases for each patient.
- EHR+LLM: This is the baseline multi-modal method that uses BlueBERT to generate clinical text embeddings. The fusing embeddings of text and structured EHR will be used to make predictions.

#### Implementation details

We adopt the data pre-processing method from CGL framework^27^. The patients with multiple visits are selected, and the last visit of each patient is used as prediction target while the history visits of each patient are used as inputs. The clinical notes “Discharge summary” from the patient will be filtered out for its high correlation with the prediction targets. The train-valid-test set are randomly split on patients with the ratio of 6000*/*125*/*1000. Each patient is used as one data sample, so one prediction will be made for each patient. We use Google T5^48^ as our LLM implementation. We use PyTorch to implement all the baseline and train/test all models on the Nvidia Tesla V100 GPU. F1-score, AUROC, and AUPRC are used as metrics to evaluate the performance of prediction. The feature importance is obtained from the attention mechanism of the predictive model.

## Results

In this section, the results of experiments aim to answer several research questions (RQ) as follows:

- **RQ1:** How does the **CKLE** framework perform in health event prediction compared to SOTA baselines?
- **RQ2:** When the labeled data is limited, can the **CKLE** still show competitive performance?
- **RQ3:** What is the contribution of each core part in the **CKLE** framework?
- **RQ4:** What are the representation learning ability of **CKLE**?

### CKLE precisely predicts the health event on multi-modal EHR data

From the Table 1, the proposed framework **CKLE** are compared with baseline methods on cardiovascular prediction tasks (Hypertension prediction and heart failure prediction). Several key findings are summarized from the results.

**Table 1.** Prediction performance on cardiovascular diseases. The number in () indicates the standard error.

| Models (Modality) | Hypertension |  |  | Heart Failure |  |  |
| --- | --- | --- | --- | --- | --- | --- |
|  | F1 (%) | AUROC (%) | AUPRC (%) | F1 (%) | AUROC (%) | AUPRC (%) |
| RETAIN (Single) | 76.02(0.33) | 75.04(0.77) | 80.24(0.78) | 68.70(0.10) | 84.53(0.06) | 76.90(0.10) |
| AdaCare (Single) | **78.89(0.42)** | 75.80(0.01) | 79.41(0.00) | 70.67(1.01) | 84.87(0.12) | 77.33(0.38) |
| Dipole (Single) | 78.13(0.98) | 73.28(2.20) | 77.82(0.68) | 70.37(0.31) | 84.40(0.62) | 76.13(0.15) |
| CGL (Multi) | 72.14(1.90) | 70.52(0.15) | 73.01(0.46) | **71.18(0.30)** | 84.79(0.20) | 73.88(1.33) |
| Chet (Single) | 77.35(0.74) | 77.77(0.11) | **80.56(0.18)** | 71.06(0.67) | 84.88(0.19) | 77.88(0.54) |
| EHR+LLM (Multi) | 77.60(0.35) | 74.79(0.29) | 76.40(1.39) | 67.47(0.76) | 84.40(0.26) | 74.03(0.91) |
| CKLE (Multi) | 78.28(0.67) | **78.14(0.86)** | 79.71(1.14) | 70.77(0.15) | **85.30(0.10)** | **78.00(0.00)** |

#### The proposed CKLE framework can surpass state-of-the-art baselines significantly on health event prediction tasks

For the hypertension prediction, we observe the **CKLE** achieves the best performance measured by AUROC. Meanwhile, **CKLE** achieves best performance on AUROC and AUPRC on the heart failure prediction. The high performance on these two cardiology related event prediction demonstrates the potential of applying CKLE for health event prediction in advance, especially for emergency medicine.

#### Modeling the patient similarity can improve the health event prediction performance

From two tasks presented in Table 1, we can observe the graph based prediction method e.g., **Chet** and the **CKLE** framework can outperform other categories of baselines including CNN-based method (Adacare), RNN-based (Diploe). The CKLE framework outperforms the RETAIN by 2.97% on F1 score and 4.13% on AUROC. The CKLE framework outperforms the Adacare by 3.01% on F1 score and 0.9% on AUROC. The reason behind this phenomenon is probably due to either graph based method or our proposed method take the patient similarity into the modeling process. We take advantage of the patient similarity to help improve the health event prediction of a particular patient. With more related information as the input, the event prediction accuracy can be reasonably increased.

#### Directly combining the text features generated from clinical text is infeasible for the performance improvement

The baseline LLM method for health event predictions directly leverages the embedded clinical text generated from LLM as the additional features don’t work efficiently to enhance the prediction accuracy. For example, compared to the backbone model, the naive LLM boosted method (indicated as EHR+LLM in Table 1) improves 2.08% F1 score on the hypertension prediction task. Meanwhile the other performance metrics on hypertension and heart failure prediction drops, which means the direct use of text features cannot improve the performance. There are two potential causes of this unsatisfying performance with additional LLM generated features. Firstly, the direct encoding of clinical text with LLM will inevitably include noise and redundant information which will affect the performance of the model. The second reason is the cross-modality knowledge distillation from LLM is more effective than naive LLM usage.

#### Cross-modality distillation from LLM is more effective than directly concatenating generated features

For the hypertension prediction, the CKLE improves the prediction performance by 3.61% on average across different baselines compared to single modality model and 4.48% on average across different baselines compared to directly using LLM generated text features. For the heart failure prediction, the average improvement is 0.709% compared to single modality model and 1.07% compared to directly using LLM. We take a further step to investigate the superiority of the cross-modality distillation strategy. The first reason is directly concatenating the features increase the dimension of the input data, which will suffer from the *curse of high dimension*. The second reason is cross-modality distillation with the proposed contrastive loss can learn the inner correlations between different modality features.

### CKLE has competitive performance with limited labeled data

To evaluate the effectiveness of **CKLE** under the limited label settings which is a common scenario in the medical application, we conduct the experiments by reducing the ratio of labeled training data on the heart failure prediction. From Figure 2, we have two observations as follows.

**Figure 2.**
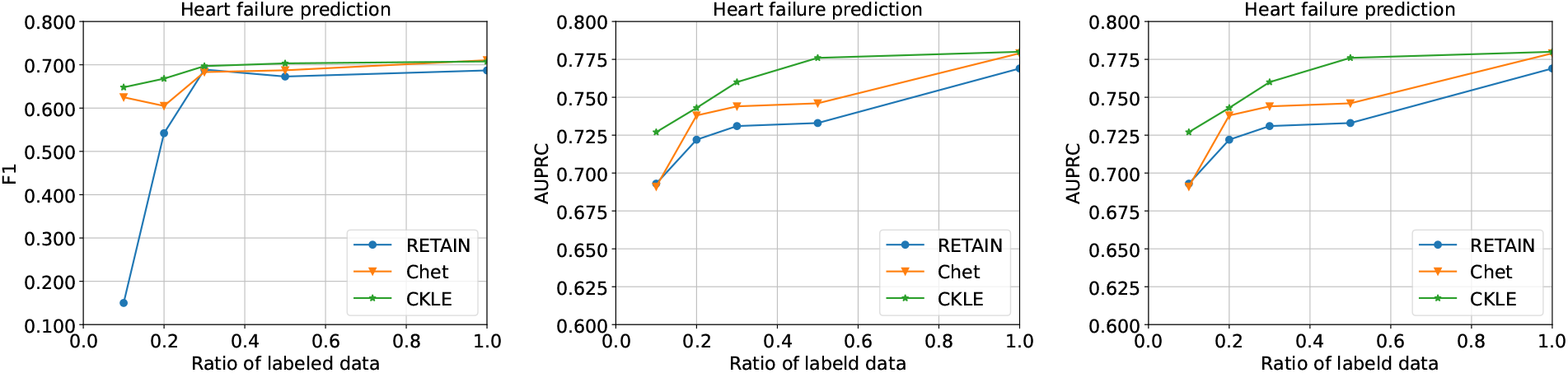
Performance comparison with limited labeled training data.

#### Increasing the amount of labeled data can increase the overall prediction performance, but the marginal effect exists

We can observe the performance of these models can gain non-trivial improvement when the ratio of labeled data is increased from 0.1 to 0.5. But the performance on whole dataset doesn’t have obvious improvement compared to half of the dataset, which indicates increasing the training data has marginal performance improvement when a threshold of enough labeled data is reached.

The CKLE framework can achieve competitive performance compared with baseline methods with limited labeled data. As presented in Figure 2, the **CKLE** framework can still surpass the baselines under different ratio of the training data. When we only use 0.1 labeled data to train the **CKLE**, we can still gain 3.18% improvement compared to the Chet model. Similarly, the **CKLE** can achieve the competitive performance with only 0.5 training data compared to the best baseline model trained on full data.

### Ablation Study

We conduct ablation study to evaluate the contribution of each part in our framework. The two key designs in the **CKLE** framework is cross-modality distillation from LLM and patient similarity modeling with contrastive loss (PSIM). The ablation study is conducted on heart failure prediction and the results are presented in Figure 3. We can observe each part has significant contribution to the performance improvement. If the knowledge is not distilled from the LLM to the predictive model, the performance is not competitive compared to the **CKLE** because the rich knowledge from LLM is powerful and helpful for various downstream health predictive tasks. Additionally, the PSIM part which leverages the patient similarity can further improve the predictive performance.

**Figure 3.**
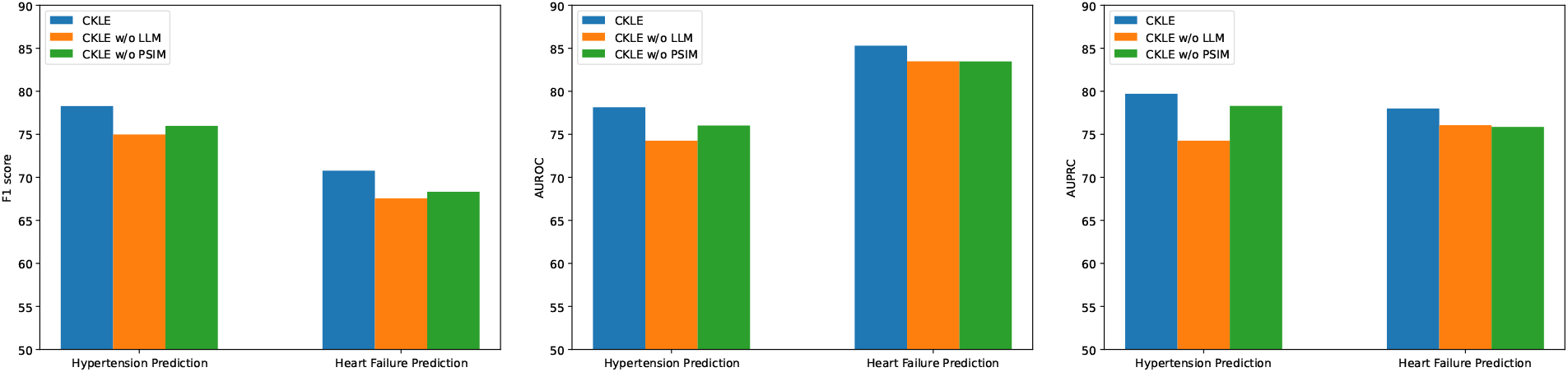
Results of ablation study.

### Embedding Visualization

To illustrate the effectiveness of the representation learning ability of the **CKLE** framework, we plot the embedding of each patients in hypertension and heart failure prediction via t-SNE. As shown in Figure 4, we compare the embeddings generated from the baseline method and the **CKLE** framework. For hypertension prediction task, the embedding visualized in Figure 4b have better clusters of negative and positive patient samples compared with the visualized embedding of the baseline method RETAIN in Figure 4a. Similarly, **CKLE** can produce better clusters of embedding on the heart failure prediction tasks.

**Figure 4.**
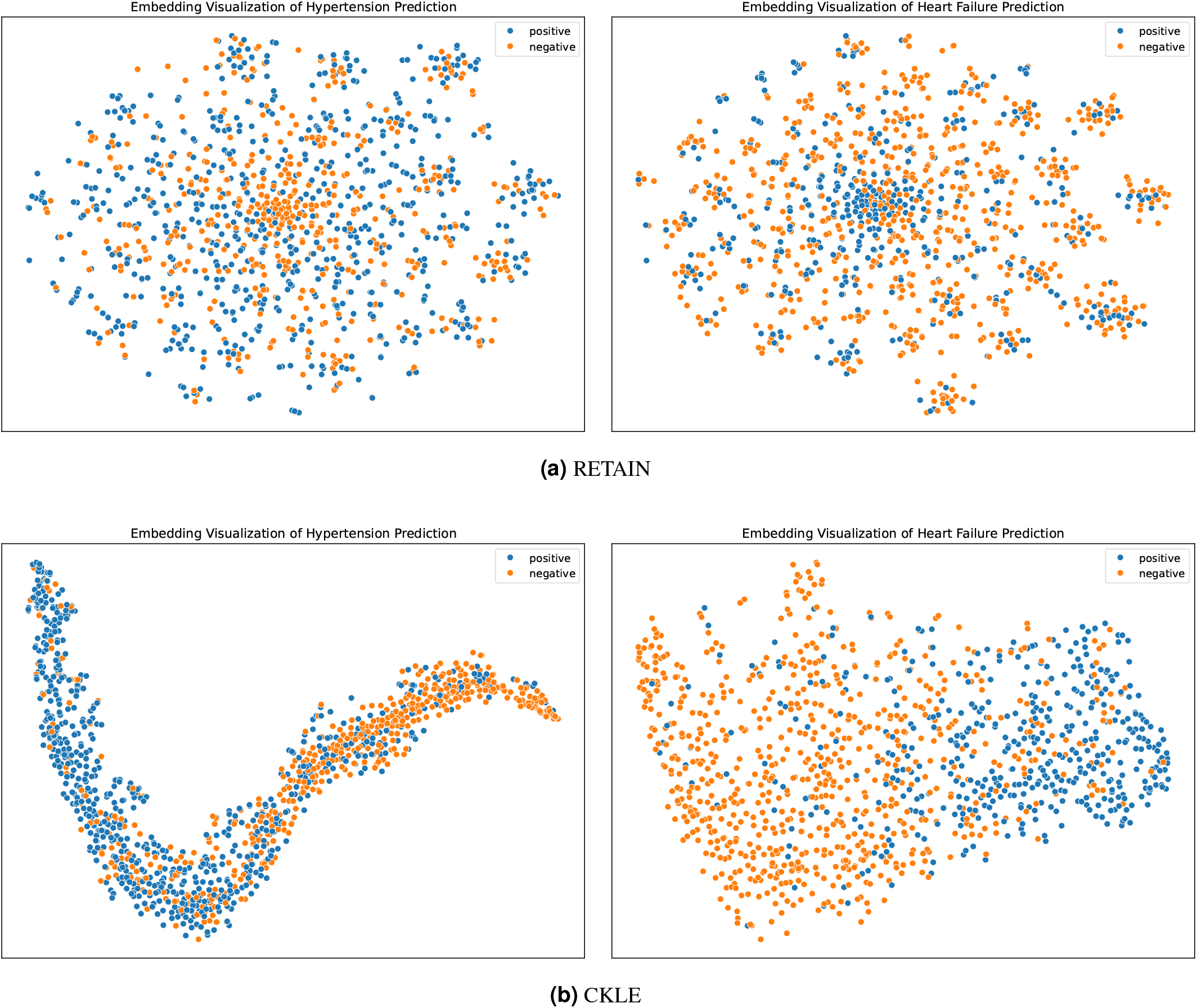
Embedding visualizations (t-SNE) on hypertension and heart failure prediction by RETAIN and CKLE.

### Model Interpretation

#### Case study I: Important Features of Hypertension Prediction

As shown in the left part of the Figure 5, we present the 20 most important features for the hypertension prediction. Each feature is assigned a score that signifies its relative importance in the prediction model. Higher scores imply a stronger relationship with the occurrence of hypertension. The most influential feature is coded 401.9, corresponding to unspecified essential hypertension, which is intuitive as it directly relates to the condition being predicted. Subsequent features include a mix of codes representing both related conditions and general health indicators. We have three observations as follows: (1). The hypertension in previous visits is an important indicator for the occurrence of hypertension the future visits. This makes sense because patients with a history of hypertension tend to be more have hypertension in future hospital visits. (2). The renal disease is highly related to the hypertension. This is a kind of complex and bidirectional relationship. The renal disease can cause and exacerbate the hypertension and vice versa. This finding also corresponds to the medical knowledge in this field^49^. (3). There are also several important features related to newborn infants, which may suggest a correlation between the circumstances of birth and the likelihood of developing hypertension later in life. Compared to hypertension in the other groups of patients, newborn hypertension is relatively rare^50^. From the salient features we observed in the hypertension prediction, the infection of infant(V29.0), respiratory problem in infant(769), feeding issue of the infant(V50.2, 779.3), preterm infants (765.19) are risk factors with high probabilities.

**Figure 5.**
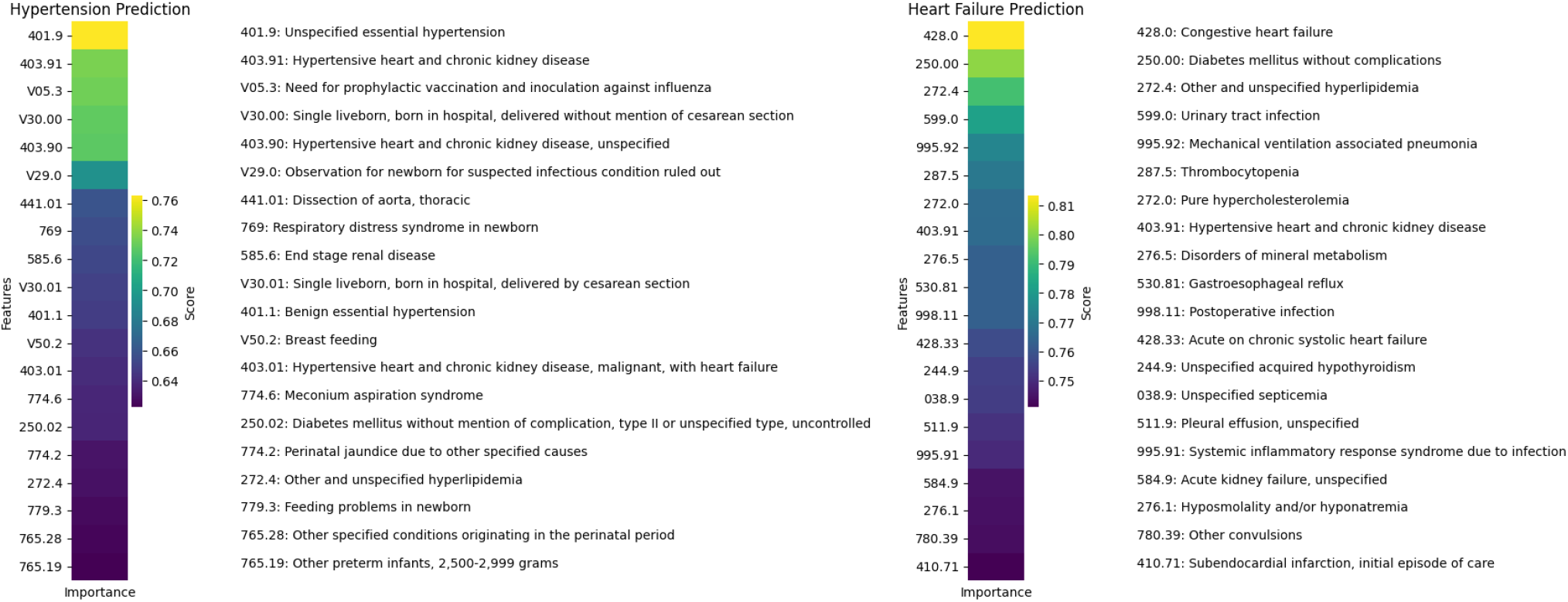
Feature importance heatmap for hypertension and heart failure prediction.

#### Case study II: Important Features of Heart Failure Prediction

In the heart failure prediction, the 20 most important features are presented on the right part of the Figure 5. There are several types of the risk factors observed from the important input features. (1). Previous cardiovascular conditions (428.0, 403.91, 428.33, 410.71) of the patients play important role in the heart failure in the future visits. (2). Heart function can also be impacted by metabolic factors (250.00, 272.4, 272.0, 276.5, 244.9, 276.1), e.g., diabetes, hyperlipidemia, and thyroid disorders. (3). Infections and postoperative complications (599.0, 995.92, 998.11, 038.9, 995.91) can exacerbate heart failure and contribute to its development or progression as well. (4). There are also other risk factors related to different organ have significant relations with the heart failure. For example, there are renal and fluid disorders (584.9, 511.9), neurological and seizure disorders (780.39), gastrointestinal disorders (530.81). Interestingly, urinary tract infection and mechanical ventilation associated pneumonia are also included, which may reflect the complex interactions between infections, treatment interventions, and heart failure risk. Hypertensive heart and chronic kidney disease and acute on chronic systolic heart failure are directly related to heart function and therefore understandably have high importance scores. Moreover, some features attract less attention from the medical field for heart failure analysis, e.g., disorders of mineral metabolism (276.5), systemic inflammatory response syndrome due to infection (995.91), postoperative infection (998.11) etc.The presence of features for subendocardial infarction and initial episode of care, underscores the multifaceted nature of heart failure risk factors and highlights the potential for machine learning models to discern complex patterns in clinical data for predictive purposes.

## Discussion

Multi-modality learning has been widely discussed and attracted lots of attention for healthcare data. The data in the healthcare domain has different characteristics compared with data in the other domains. From the standpoint of data-centric AI, three healthcare data challenges are summarized as follows: (1). The noise in the healthcare data is prevalent and unignorable. This can be caused by the device noise, human bias, noise in recording process, etc. (2). The clinical text is usually highly specialized and domain specific. There are lots of professional terms in the medical domain, which requires prior knowledge. (3). There is usually a privacy issue of the health data to request the model deployed locally. Most hospitals will not put their data on the cloud server or use the online models to help with their clinical workflows. These data level challenges put out the requests to design novel and suitable AI models with an emphasis on the precision, robustness and privacy.

Previous works mainly advance the technique and models to learn the embedding of different modalities and combine them in an efficient way. In the realm of LLM, the representation learning ability from text data has been transformativly boosted. How to efficiently leverage the LLM in the multi-modality healthcare data remains as an open research question. We distinguish the **CKLE** framework from the related work from four aspects. (1) The knowledge of LLM is effectively learned by the health predictive model with knowledge distillation. The knowledge from LLM is powerful which leads to large-scale parameters of the LLM that has efficiency issue. (2). We explore a novel method of patient similarity learning with contrastive loss function. The patient similarity can be learned by taking advantage of the contrastive loss, which can be used to learn postive and negative pairs. We design the soft labels for the contrastive loss function to learn the similarities between patients with more granularity. This contrastive loss for patient similarity learning can be easily adapted to other predictive model by inserting into the loss function. (3). Besides competitive prediction accuracy, the **CKLE** framework can learn better representations validated by embedding visualization. From the observations in the Table 1, the increase of performance metrics indicates the effectiveness of the **CKLE** to improve the predict accuracy. However, we cannot observe the representation effectiveness through the numerical results, which are also very important to evaluate the model. In the t-SNE embedding visualization experiments, we can observe a more clear discrimination between two categories predicted by the **CKLE** compared to the baseline method. (4). **CKLE** predictive model preserves the global model interpretability, which can provide the feature importance by the attention score. The interpretability is a very essential aspect when we build the medical AI models. In this paper, we distill the knowledge from LLM into the predictive model, which is a type of Transformer. The interpretability of Transformer can be represented as the attention score for each input features. The feature importance can show which feature plays an important role in the predictions. From the model interpretation analysis, we study two cases on hypertension and heart failure prediction. The top 20 important features we get corresponds to the medical knowledge with the domain expert. So our model can produce precise as well as interpretable predictions on the health events.

From the collaboration with domain expert in the cardiology diseases, we can validate some already known medical knowledge and discover some new features which lacks enough attention previously. The **CKLE** can not only precisely predict health events but also can discover some medical findings.

## Data Availability

All data produced are available online at MIMIC.

https://physionet.org/content/mimiciii/1.4/

## Data Availability

The data that support the findings of this study are available from PhysioNet but restrictions apply to the availability of these data, which were used under license for the current study, and so are not publicly available. Data are however available from the authors upon reasonable request and with permission of PhysioNet.

